# The phenomenology of auditory verbal hallucinations in schizophrenia assessed with the MiniVoiceQuestionnaire (MVQ)

**DOI:** 10.1101/2023.02.16.23285636

**Authors:** Kenneth Hugdahl, Helene Hjelmervik, Sarah Weber, Lydia Brunvoll Sandøy, Josef Bless, Lin Lilleskare, Alexander Craven, Marco Hirnstein, Katarzyna Kazimierczak, Gerard Dwyer, Magda L. Dumitru, Igne Sinceviciute, Lars Ersland, Erik Johnsen

## Abstract

We used a 10-question self-report questionnaire, Mini Voice Questionnaire (MVQ), for mapping the phenomenology of auditory verbal hallucinations (AVH). The MVQ contains questions related to daily AVH frequency and duration, the events preceding AVH episode onset and offset, the very first AVH episode, emotional content, coping strategies, if the voice comes from the inside or outside of head, if it is one’s own voice heard, and whether the voice is present when filling out the questionnaire. Forty-one patients with a diagnosis of schizophrenia spectrum disorder participated in the study. The construction of the MVQ was originally driven by an interest in whether AVH-episode onsets and offsets, that is, the coming and going of the voice, are initiated by specific environmental events or mental states, or whether they occur spontaneously, which could have both theoretical and clinical implications. MVQ scores were correlated with PANSS and BAVQ questionnaire scores. The results showed that specific events do not precede onset or offset of AVH episodes except for the very first episode which was often associated with trauma or other negative events. This finding could have implications for neurobiological models of AVH, showing that AVH episodes are spontaneously initiated, pointing to a neuronal origin of AVH episode onsets *and* offsets. The P3 (hallucinatory behavior) item of the PANSS questionnaire correlated significantly with frequency and duration of AVH episodes: More frequent and longer AVH episodes were associated with higher P3 scores, implying more severe symptoms. The results are discussed in terms of recent AVH models.

## Introduction

The phenomenology of AVH is typically studied with questionnaires, and the most widely used are the Positive and Negative Syndrome Scale (PANSS) (Kay et al., 1987), the Psychotic Symptom Rating Scales (PSYRATS) (Haddock et al., 1999), the Auditory Vocal Hallucination Rating Scale (AVHRS) (Jenner & Van de Willige, 2002), and more recently the Questionnaire for Psychotic Experiences (QPE) (Rossell et al., 2019). A somewhat different, but frequently used scale is the Beliefs About Voices Questionnaire (BAVQ) (Chadwick et al., 1995) which is focused on the emotional content of the voices. The structure of all these questionnaires varies, and some are initiated by a clinician while others are self-reports, but a common theme is that they are broad scales (perhaps with the exception of AVHRS), covering additional symptoms and processes. There are however exceptions also here, the QPE for example makes a distinction between AVH and other sensory modalities. The PANSS, perhaps the most widely used scale, has only one item for hallucinations, of a total of 30 items, and even this item is not specific for auditory hallucinations, although AVHs are emphasized.

A feature of AVHs, typically not covered by traditional questionnaires, is the seemingly fluctuating nature of hallucinatory episodes, that few, if any, patients have episodes that are constantly present (see Hugdahl, 2015 for a discussion). AVHs fluctuate over the course of a day, despite great individual variation in frequency and duration of episodes (Bless et al., 2020). Research on the phenomenology of AVHs has primarily focused on understanding the onset, or elicitation, of an episode, with little or no focus on the offset or inhibition of an episode (but see Hugdahl et al., 2022).

Coupled with this is the question of whether such day-to-day fluctuations are environmental or spontaneous without an apparent trigger. In other words, are the onset and offset of hallucinations controlled from the out- or inside of the head? An answer to this question could have both theoretical and practical implications. Theoretically, if onsets and offsets are triggered from the inside, research could be directed to finding the underlying neurobiology of such spontaneous triggers, and new treatments could be developed to prolong offsets or even inhibit new onsets once it is discovered what causes the offset in the first place. There is probably a long way to reach such a level of understanding, but a first small step could be to collect data on the phenomenology of AVH-episode onsets and offsets, addressing the question of whether hallucinating individuals experience that fluctuating AVH episodes are linked to specific environmental events, and if so what events? If AVH episodes are triggered from the outside, effective treatments could simply try to prevent such triggering stimuli.

Since AVH is closely linked to a schizophrenia diagnosis, the initial outbreak of an AVH episode will therefore be correspondingly linked to the onset of schizophrenia (Larøi et al., 2012). Traumatic experiences such as child and adolescent violence and sexual abuse are precursors of AVH (Nesbit et al., 2022; Misiak et al., 2016), although the mediating pathways are not entirely clear. Similarly, illicit drug use, in particular cannabis, has been suggested to mediate the initial onset of AVHs (Andrade et al., 2016; Matheson et al., 2022), coinciding with psychosis onset. Distressing life-events therefore seem to be mediators of both psychosis and positive symptoms, like AVHs (Næss et al., 2022).

An interesting question is however the degree to which recurring episodes later in life, after a diagnosis, also are linked to phenomenologically distressing events, or if they occur in the absence of such events. To address the question of to what degree environmental events or internal states precede the onset and offset of an episode, we constructed a simple questionnaire, the Mini Voice Questionnaire (MVQ).

In addition, we also wanted to tap other AVH dimensions such as episode frequency and duration, preceding events before the onset of the very first episode, emotional content of the voice, if the voice is experienced as coming from inside or outside of the head, strategies to mentally cope with the voice, whether it is one’s own or somebody else’s voice, and whether the voice is present or not at the time when filling out the questionnaire.

We collected MVQ data from 41 patients with a schizophrenia diagnosis as part of a larger study on the psychology and neurobiology of AVH (https://erc2group.w.uib.no/). The MVQ data were used in conjunction with other questionnaire data such as PANSS and BAVQ, and correlations with the data from these questionnaires are presented, particularly correlations with the PANSS P3 item which taps severity of hallucinations.

## Method

### Participants

The participants were 41 patients who all experienced AVH; 26 had an ICD-10 diagnosis of paranoid schizophrenia (F20), 5 with schizophrenia unspecified non-organic psychosis (F29), 3 with schizoaffective disorder (F25), 2 with drug-induced psychosis (F19), 2 with personality changes (F62), 1 with bipolar disorder (F31), and 2 with no formal diagnosis. There were 15 males and 26 females in the sample, one of the participants self-reported to be male, although born as female.

All participants, except six, were on antipsychotic medication, mostly second-generation antipsychotics. Medication was defined and calculated as Defined Daily Doses (DDD) according to WHO https://www.who.int/tools/atc-ddd-toolkit/about-ddd (see also Leucht et al., 2016). The definition of DDD is the assumed average maintenance dose per day for a drug used for its main indication in adults. This procedure showed that mean DDD for the whole group was 0.98. The majority of the patients (71%) were on either Clozapine, Quetiapine or Aripiprazole either as mono-therapy or in combination with other drugs, prescribed either as oral or depot.

The patients were recruited in the Bergen City through the Vestland County Health Care System, primarily from the Sandviken Psychiatric Clinic, Haukeland University Hospital (HUH) in Bergen, Norway. Knowledge of the project was made available to psychiatric health-care personnel through announcements on boards and through social media, as well as through distributed brochures and leaflets. A few patients were recruited from psychiatric wards from other counties in Western Norway. Brochures and other kinds of information were in addition placed at regional and local psychiatric units, with invitation to health- personnel to contact the project nurse if the project seemed relevant for her. The project nurse also participated in weekly staff meetings at Sandviken Psychiatric Clinic HUH and at other psychiatric outpatient clinics in the Bergen city area.

### MiniVoiceQuestionnaire (MVQ)

The 10 questions making up the MVQ (see Table 1) dealt with frequency of AVH episodes (#1 VoiceFrequency); duration of an episode (#2 VoiceDuration); events preceding the onset of an episode (#3 VoiceStart), events preceding the offset of an episode (#4 VoiceStop); events preceding the first experience of an episode (#5 VoiceFirst); negative emotional content of episodes (#6 VoiceContent); location of episodes, inside or outside head (#7 VoiceHead); behavior to avoid or escape episodes (#8 VoiceCoping); one’s own or other’s voice (#9 VoiceOwn); voice present at the time of filling out questionnaire (#10 VoicePresence). The response alternatives for each question were bimodal for eight questions (“yes” or “no”), with the first two questions (frequency and duration) having three response alternatives (see Table 1). For AVH frequency these were 1-3 episodes/day, 4-5 episodes/day,>5 episodes/day. For AVH duration the corresponding alternatives were <10 min/day, 10-30 min/day, and >30 min/day.

**Table 1:** The MVQ questionnaire. Copies in pdf-format can be obtained from Kenneth Hugdahl

|  |  |
| --- | --- |
| **<sup>1</sup>ERC2 Mini Voice Questionnaire (MVQ)**<br>ID:<br>DATE:<br>1. How many times per day do you normally hear the voice(s)? **(VoiceFrequency)**<br>1-3<br>☐<br>3-5<br>☐<br>5 <<br>☐<br>2. For how long do you normally hear the voice(s) each time? **(VoiceDuration)**<br>< 10 min<br>☐<br>10-30 min<br>☐<br>30 min <<br>☐<br>3. Is there anything in particular that normally happens just before you hear the voice(s) in everyday life? **(VoiceStart)**<br>☐ Yes ☐ No<br>What happens?<br>4. Is there anything in particular that normally happens just before the voice(s) stop in everyday life? **(VoiceStop)**<br>☐ Yes ☐ No<br>What happens?<br>5. Do you remember if anything special happened the first time you heard the voice(s)? **(VoiceFirst)**<br>☐ Yes ☐ No<br>What happened? | 6. Would you say that the voice(s) is mostly evil/ill-natured? **(VoiceContent)**<br>☐ Yes ☐ No<br>What does the voice(s) say?<br>7. Does the voice(s) normally come from outside of your head? **(VoiceHead)**<br>☐ Yes ☐ No<br>8. Do you normally do anything in particular to avoid or escape the voice(s)? **(VoiceCoping)**<br>☐ Yes ☐ No<br>What do you do?<br>9. Is it your own voice that you hear? **(VoiceOwn)**<br>☐ Yes ☐ No<br>10. Do you hear the voice(s) right now? **(VoicePresence)**<br>☐ Yes ☐ No |

<sup>1</sup> <sup>1</sup> Kenneth Hugdahl, IBMP, Universitetet i Bergen, og Divisjon for psykisk helsevern, Haukeland Universitetssykehus, Bergen

Differences in the distribution of “yes” and “no” scores for the eight bimodal questions in the MVQ questionnaire was tested for statistical significance with the Binomial test (SPSS v.27 https://www.ibm.com/products/spss-statistics) under the assumption of equal (50/50%) probabilities for the “yes” and “no” scores. The AVH Frequency and Duration question scores were tested under the assumption of 33/33/33% equal probabilities for the three alternatives, using the One-sample Chi^2^ test in the SPSS package.

We also collected narratives of what the participants subjectively commented when filling out the MVQ questionnaire, for example if a participant indicated a “yes” to the VoiceStart question, a follow-up question would be “What happened?”, and they then wrote down their answers.

### PANSS questionnaire

The PANSS is an interview questionnaire (Kay et al., 1987) which covers positive, negative, and general psychopathology symptoms. Each symptom contains questions that the clinician asks the patient and then evaluate the answer into a composite symptom score ranging from 1 (no symptom load) to 7 (extreme symptom load). In the present analyses, the PANSS P3 (Hallucinatory behavior) and the sum of PANSS Positive and sum of PANSS Negative symptoms were used.

### BAVQ questionnaire

The BAVQ-R scale is a self-report scale with several different sub-scales, that contains a set of statements that the respondent answers on a scale from 0 (Disagree) to 3 (Agree strongly) (Chadwick et al., 2000). The sub-scales included in the present analyses were Malevolence (M) with statements about negative emotional content of the voice, like “My voice is evil”, Benevolence (B) with statements about positive emotional content of the voice, like “My voice wants to help me”, and Omnipotence (O) with statements like “My voice is very powerful”. Each of these sub-scales contain six statements, thus the maximum score for a sub- scale is 18. Scores for each sub-scale were summed and averaged for each participant and the mean value was entered into the analyses. The PANSS and BAVQ scores were correlated with the MVQ scores using the Spearman statistic for non-parametric scores.

## Results

The distribution of scores is seen in Figure 1 as bar graphs with sample size for each question and with corresponding percentages added to the bars in the graphs.

**Figure 1:**
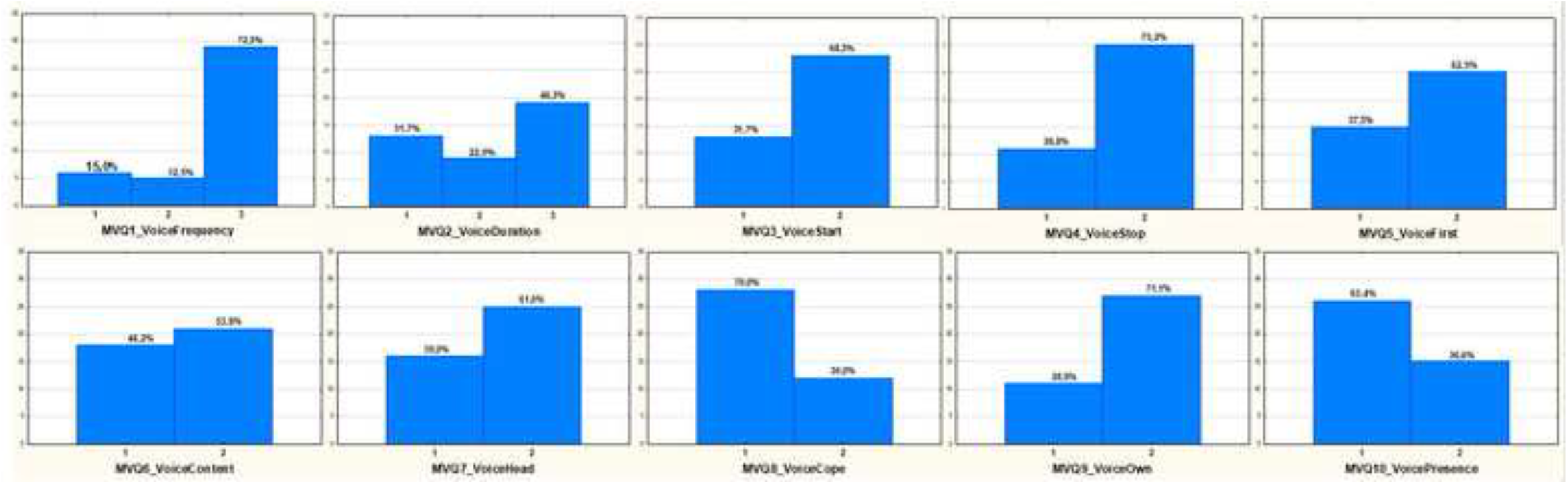
Bar graphs showing the distribution of individuals answering “Yes” or “No” to the eight questions with a bimodal and the two questions with a trimodal response structure in the MVQ. The Y-axis show absolute numbers, with corresponding % marked above respective bar. MVQ1_VoiceFrequency: 1 = “1-3 times/day”, 2 = “3-5 times/day”, 3 = “More than 5/times/day”. MVQ2_VoiceDuration: 1 = “<10 min/day”, 2 = “10-30 min/day”, 3 = “More than 30 min/day”. For all other MVQ questions: 1 = “Yes”, 2 = “No”.

Figure 1 shows that significantly more participants answered “No” (68,3%) compared to 31.7% who answered “Yes” to VoiceStart question. The difference was significant (*p* = .029). The same was true for the VoiceStop question, where 73.2% answered “No” and 28.8% answered “Yes”, (*p* = .005). Overall, significant differences were also found for the VoiceFrequency (*p* = .001), VoiceCope (*p* = .018) and VoiceOwn (*p* = .015) questions. These differences showed that significantly more participants reported experiencing more than 5 episodes per day, 72.5% versus 15.0 and 12.5 % (VoiceFrequency), that more patients tried different coping strategies when experiencing AVH episodes than those that did not try coping strategies, 70.0% versus 30.0% (VoiceCope), and that the majority did not experience their own voice during episodes, 71.1% versus 28.9% (VoiceOwn) (see Figure 1 for further details). Considering alpha inflation with critical alpha (*p*) set to .05 one would expect that 1 out of 20 tests (5%) would be significant by chance. The present result of five out of 10 tests being significant (50%) exceeds what would be expected by chance.

The following questions were non-significant: VoiceDuration, VoiceFirst, VoiceContent, VoiceHead, and VoicePresent, although as seen in Figure 1, there was a trend for more “No” answers compared to “Yes” answers for VoiceFirst, VoiceContent, and VoiceHead, indicating that more participants did not recall specific events preceding the first AVH episode (62.8% versus 37.8%), reported that the voice more often (61.0%) came from the inside compared to the outside (39.0%) of the head (VoiceHead), and experienced AVH when filling out the MVQ questionnaire (63,4%) compared to (36.6%) “No” responses (VoicePresence

### Subjective comments

The following are examples of what the participants said to the follow-up questions. Those who reported events preceding the onset of episodes (VoiceStart) often said that AVH episodes followed *“when I have periods of anxiety and emotionality”, “when I experience prolonged stress periods”, “when I have problems sleeping”, “when I am uncertain of what is happening around me”*. For the VoiceStop question, participants said that the voice tended to “*disappear when I manage to distract myself”; “when I shift attention focus away from the voice”; “when I am in a better mood”, or “when the medication work”*. As for the VoiceFirst question, those who reported events preceding the onset of the voice said that it could happen when *“I have bad consciousness for not having stood up for myself”; “when I am stressed and exhausted”; “when I worry about things”; “when someone is listening to my thoughts”, or “when I think a lot about life”*. For the VoiceContent question, those participants who had indicated that the content was primarily negative said that the voice demanded that *“I should hurt myself”; “I should shut up, and that I am stupid”; “I hear the voice of Satan”, “you should not be here”, “the voice gives me bad conscience”, “the voice is commanding me”* as examples. Examples of voices with positive content were “*I hear singing”, “I hear the voice of God who tells me it will be OK”*. As for coping strategies (VoiceCoping), the participants provided a mixed activities that they tried, like *“When I start doing something”, “I try to divert myself away from the voice”, “I listen to music”, “I watch movies”, “I try to relax or go to sleep”*.

### PANSS and BAVQ scores and correlations with MVQ scores

The mean PANSS P3, mean sum PANSS Positive and mean sum PANSS Negative symptom scores were respectively 4.65; 18.77, and 18.02. The corresponding means for the BAVQ Malevolent (M), Omnipotent (O), and Benevolent (B) sub-scales were 15.94, 15.09, and 11.34, respectively. A One-way ANOVA and follow-up tests with Tukey’s HSD-test for the BAVQ statements revealed that the scores for the Malevolent (M) and Omnipotent (O) statements were significantly higher than the scores for the Benevolent (B) statements (*p* =.006 and *p* = .020, respectively), with no significant difference between the M and B statements (*p* = n.s.). There was no significant difference between sum PANSS Positive and sum Negative scores (*p* = n.s.)

Table 2 shows correlations between MVQ scores on the one hand, and PANSS P3, PANSS Positive, PANSS Negative scores, BAVQ Malevolent (M), Benevolent (B), and Omnipotent (O) scores, on the other hand.

**Table 2:** Spearman rank-order correlation coefficients between the 10 MVQ questions and PANSS P3 and the separate sums of PANSS Positive and Negative scores and mean BAVQ Malevolent (M), Benevolent (B) and Omnipotent (O) scores. Significant (*p* <.05) correlations are marked in red, near-significant (*p* ≤.060) are marked in orange.

|  | PANSS_P3 | PANSS Pos | PANSS Neg | BAVQ_M | BAVQ_O | BAVQ_B |
| --- | --- | --- | --- | --- | --- | --- |
| **VoiceFreq** | **0,624** | **0,295** | **0,310** | 0,007 | -0,048 | **-0,337** |
| **VoiceDur** | **0,360** | 0,054 | 0,126 | -0,062 | -0,181 | 0,022 |
| **VoiceStart** | 0,075 | 0,134 | **0,377** | -0,078 | -0,116 | -0,086 |
| **VoiceStop** | -0,043 | 0,083 | 0,160 | 0,158 | 0,026 | -0,286 |
| **VoiceFirst** | -0,130 | -0,153 | 0,115 | 0,271 | 0,096 | 0,028 |
| **VoiceCont** | -0,250 | -0,010 | -0,005 | 0,111 | 0,181 | -0,071 |
| **VoiceHead** | **-0,296** | -0,199 | -0,122 | 0,162 | 0,127 | -0,066 |
| **VoiceCope** | -0,063 | 0,087 | **0,339** | -0,031 | 0,038 | 0,019 |
| **VoiceOwn** | 0,191 | 0,217 | 0,208 | 0,210 | -0,036 | -0,165 |
| **VoicePres** | **-0,330** | -0,020 | -0,134 | -0,148 | -0,051 | 0,125 |

MVQ VoiceFrequency and VoiceDuration scores correlated positively with PANSS P3 scores, meaning that the more frequent and with longer duration AVH episodes were, the more severe were they evaluated (*p* = .00002 and .021, respectively). There was also a significant correlation for VoiceStart and PANSS Negative scores (*p* = .016). There was a significant negative correlation between VoicePresence and PANSS P3 (*p* = .035), indicating a relationship between presence of AVH and PANSS P3 scores. Finally, there was a significant positive correlation between VoiceCope and PANSS Negative symptoms (p =.035) indicating that more negative symptoms were associated with less coping behavior.

In addition, there were tendencies for the correlations between VoiceFrequency on the one hand and PANSS Positive (*p* = .069) and PANSS Negative scores (*p* = .055) on the other hand. Thus, AVH episodes that were more frequent and with longer duration were associated with increased hallucinatory severity. Other correlations approaching significance were VoiceHead and PANSS P3 (*p* = -.060) and VoiceFrequency and BAVQ Benevolence (B) (*p* =.051). The first of these correlations indicated that voices experienced as coming from the outside of the head were associated with increased AVH severity, while the second correlation indicated that the more frequent AVH episodes, the less benevolent were the voice experienced. With 60 correlations and a *p*-value threshold of .05, three correlations would appear by chance (5/100 × 60).

### Sex differences

Effects of the sex factor were statistically tested with 2 × 2 Chi^2^ for the VoiceContent, VoiceDuration, VoiceFirst and VoicePresence MVQ questions. This showed non-significant effects for comparisons involving sex as a factor. It could be mentioned though that more males answered “No” to the question if the voices were predominantly negative (15/10) while more females answered “Yes” to the same question (8/6). As for the PANSS and BAVQ scores, there were no significant sex differences when tested with independent sample t-tests (all *p* > .05).

## Discussion

Regarding triggering events for AVH episode onset (VoiceStart) and offset (VoiceStop), that is, fluctuations of AVHs, the results clearly showed that the majority of the participants did not recall or recognize such preceding events. This was different from the first recalled episode (VoiceFirst) which was associated with negative events, like trauma, stress, and drug abuse, although the difference was non-significant. Other significant, or near-significant, MVQ score distribution differences were for VoiceFrequency, VoiceCope and VoiceOwn, indicating that a majority of the participants experienced more frequent (also with longer duration) AVH episodes, which also correlated with severity of AVHs, that the voice was not their own, and that it came from outside the head, and that a majority tried various coping strategies, like listening to music and diverting themselves from the voices, which also correlated with PANSS negative scores. The finding that episode onsets and in particular, offsets seem to occur spontaneously, in the absence of eliciting environmental events or inner mental states, could have both theoretical and clinical implications. Voice frequency and duration

The results resonate with the recent finding by Hugdahl et al. (2022) that onsets and offsets have different underlying neuronal mechanisms. Hugdahl et al. (2022) used fMRI to track neuronal events second-by-second for 10 seconds before to 15 seconds after patients indicated the start or stop of an AVH episode by pressing a hand-held button. Results of this study showed that an activation response occurred 5 seconds before the patient indicated being aware of the presence of the voice, and a similar response being initiated 2 seconds before the patient became aware of the disappearance of the voice (see also Shergill et al., 2004 and Hoffman et al, 2008 reporting similar findings). Relating the present MVQ findings to the imaging findings of Hugdahl et al. (2022) implies that AVH episodes have an endogenous cause, that is, they appear spontaneously and independent of exogenous events, not ruling out the possibility that such events could have influenced subconsciously. In general, the answers to the VoiceStart and VoiceStop questions were consistent with the answer to the VoiceFirst questions, that is, if the answer to the VoiceFirst question was “no”, the most likely answer to VoiceStart and VoiceStop questions would also be “no”, and vice versa.

Interestingly, since not all participants reported absence of precipitating events, those that did report such events mostly reported psychological distressing situations at the onset of an episode, similar to what Larøi (2012) reported. The opposite was evident for participants who reported specific situations preceding the offset of an episode. As for correlations, the strongest associations were for frequency and duration of episodes and severity of AVHs as judged from the PANSS P3 item. Although PANSS P3 covers a follow-up question of “how often you hear these voices or messages”, a PANSS P3 score is a measure of overall severity. The present finding of a significant positive relationship between severity on the one hand and frequency and duration on the other hand is therefore a demonstration of a direct association between how often AVH episodes are experienced and how distressing and severe they are.

That patients who experience frequent AVH episodes engage in various coping strategies is a known phenomenon, typically in the form of distracting activities to shift focus away from the inner voices to the outside world. This has also been the target for cognitive behavioral treatment attempts to enhance or boost coping with AVHs (Hayward et al., 2022; Wiersma et al., 2001). The present finding of a significant correlation between MVQ VoiceCope and PANSS Total Negative score therefore adds to the positive effect of applying various coping strategies, not only for positive but also for negative symptoms (cf. Hayashi et al., 2007).

The near-significant negative correlation between VoiceHead and PANSS P3 implies that the more severe AVHs are experienced the more often the voice appears as coming from the outside of the head, which substantiates previous findings of a similar relationship. For example, Lera et al. (2011) found that patients who experienced the voice outside of the head reported less insight into their condition, which the authors interpreted as that these patients lacked understanding that their voices were produced by their own mind.

The question on emotional quality of the voice (VoiceContent) gave a somewhat unexpected result, where negative emotional content did not differ from non-negative content. This was surprising considering the often reported finding that about 80% of a schizophrenia spectrum diagnosis report negative voice content (de Leede-Smith et al., 2013; Daalman et al., 2011; Bless et al., 2018; Larøi, 2012). It could have been caused by the way this question was framed, *“Would you say that the voice(s) is mostly evil/ill-natured”*, which may have emphasized strong malevolence, thus ignoring more subtle aspects of negative emotionality. This is substantiated by the finding for the BAVQ statements with significantly higher scores for BAVQ M and O statements compared to B statements. It should perhaps also be mentioned in this context that de Boer et al (2022) in a recent paper found that only 34.7% of their clinical participants reported negative emotional content, although this was higher than the percentage of reports in a non-clinical control group (18.4%). It should also be noted that negative emotional content is not always associated with aversive life-experiences. For example, Daalman et al (2012) found no difference in emotional voice content between a group of AVH individuals who had a history of abuse and trauma compared with a group without such experiences (see also Næss et al., 2022).

The current results for VoiceCoping are in line with previous findings that patients try various coping strategies when voices are experienced as negative, often some kind of diversion strategies, to avoid the negative impact of the voices (e.g. Hayward et al, 2022).

There was also a significant effect of VoiceOwn, where a majority of the participants answered that they did not experience hearing their own voice (71% versus 29%). This finding could add to the understanding of the underlying phenomenology, in particular models of AVHs as caused by disturbed monitoring of inner speech (McGuire et al., 1995), or as inner speech being misinterpreted as coming from outer sources (Heinks-Maldonado et al., 2007). Ever since its inception as a model for understanding the phenomenology of AVH, the “inner speech model” has been met with both supporting and opposing empirical evidence, including also studies to establish a neuronal basis for the model. In a recent review of 417 papers, Barber et al. (2021) concluded that *“…evidence was found to both support and oppose the inner speech model”*. A common definition of inner speech in the literature was given by Levine et al. (1982), who defined inner speech as “*subjective phenomenon of talking to one self”*, see also Jones & Fernyhough (2007) for a critical discussion of the concept of inner speech. An implication of such a definition would be that the individual would be hearing one’s own voice, which the current finding clearly did not support.

## Data Availability

Due to clinical data regulations imposed by the Western Norway Ethical Committee (REK-Vest) (https://rekportalen.no/#hjem/home), data can be shared by request to Kenneth Hugdahl, and after written permission from the REK-Vest.

## Funding and ethical approval

The research reported in this paper was funded by an Advanced Grant from the European Research Council (ERC) #693124 and had ethical clearance from the ERCEA (#693124) and the Regional Committee for Medical Research Ethics in Western Norway (REK Vest #2106/800). Due to clinical data regulations imposed by the Western Norway Ethical Committee (REK-Vest) (https://rekportalen.no/#hjem/home), data can be shared by request to Kenneth Hugdahl, and after written permission from the REK-Vest.

## Contributors

**Kenneth** Hugdahl, designed the study, analyzed data, wrote the ms

**Helene Hjelmervik**, discussed the design and analysis of data, commented on the ms

**Sarah Weber**, discussed the design and analysis of data, commented on the ms

**Lydia Brunvoll Sandøy**, helped collect data, assisted in data analysis, commented on ms

**Josef Bless**, took part in design of study, discussed and commented on the ms

**Lin Lilleskare**, collected data, took part in discussions of data analysis

**Alexander Craven**, discussed the design and analysis of data, commented on the ms

**Marco Hirnstein**, discussed the design and analysis of data, commented on the ms

**Katarzyna Kazimierczak**, took part in data analysis, discussed analysis of data

**Gerard Dwyer**, discussed the design and analysis of data, commented on the ms

**Magda L. Dumitru**, discussed the design and analysis of data, commented on the ms

**Igne Sinceviciute**, took part in data collection, commented on the ms

**Lars Ersland**, discussed the design and analysis of data, commented on the ms

**Erik Johnsen**, took part in data collection, commented on the ms

## Conflict of interest

The authors declare no conflict of interest.

## Role of the funding source

The research presented in the submitted ms was funded by a grant to the first, corresponding, author, Kenneth Hugdahl from the European Research Council (ERC), #249516 and from the Western Norway Health Authority (Helse-Vest) #912045. The funding sources provided economic support for the research.

## Acknowledgment

The authors would like to acknowledge the contribution of patients and their families for participating in the study.

## Notes

### Competing Interest Statement

The authors have declared no competing interest.

### Funding Statement

The research reported in this paper was funded by an Advanced Grant from the European Research Council (ERC) #693124, and from the Western Norway Health Authorities Grant #912045

### Author Declarations

Ethical clearance was provided from the European Research Council Executive Agency (ERCEA #693124) and from the Regional Committee for Medical Research Ethics in Western Norway (REK-Vest #2106/800)

